# Epidemiology of antimicrobial use in Malawi: a cross-sectional study using World Health Organisation core antimicrobial use indicators in faith-based health facilities

**DOI:** 10.64898/2026.07.01.26357065

**Authors:** Evans Chimayi Chirambo, Francis Kachidza Chiumia, Dumisani Enricho Nkhoma, Collins Mitambo, Alex Thawani, Taonga Msiska, Sharon Odeo, Judith Asin, Matias John, Waratchaya Chuaikan, Martin Angwe, Patience Khomani, Innocent Chibwe, Simon Matchado, Chikhulupiliro Chimwaza, Pacharo Matchere, Beatrice Chiweza, Elled Mwenyekonde, Elizabeth Kampira, Elimase Kamanga, Zione Salima, Clifford George Banda, Happy Makala

## Abstract

**Background:** Antimicrobial resistance (AMR) is a major public health threat globally, with a disproportionate burden in sub-Saharan Africa. Faith-based health facilities provide essential healthcare services to underserved populations, yet data on antimicrobial use in these settings remain limited.

**Aim:** To assess antimicrobial use in Christian Health Association of Malawi health facilities using the World Health Organisation core medicine use indicators.

**Materials and Methods:** A multicentre cross-sectional study was conducted in 29 CHAM health facilities across Malawi between January 2024 and June 2025. Data were collected from facility personnel, inpatient prescriptions, and patient interviews and analysed using descriptive and inferential statistics.

**Results:** Average availability of key antimicrobials was 33.1% (95% CI: 29.7–36.4), while customised formularies were available in 64.3% of health facilities. Among 660 prescriptions analysed, 90.3% contained an antimicrobial agent, but only 33.2% adhered to standard treatment guidelines and 43.6% were prescribed using full generic names. Facilities with pharmacy professionals were more likely to have a facility-specific formulary (84.6% vs. 46.7%, p = 0.037).

**Conclusion:** Antimicrobial stewardship gaps remain substantial in faith-based health facilities in Malawi and across sub-Saharan Africa, highlighting the need for targeted stewardship programmes in faith-based health facilities.

## Introduction

Antimicrobial resistance (AMR) occurs when microorganisms develop the ability to survive exposure to antimicrobial agents that would normally inhibit or eliminate them at established therapeutic doses. AMR has emerged as one of the greatest threats to global public health, compromising the prevention and treatment of infectious diseases and threatening decades of progress in modern medicine. The World Health Organisation (WHO) estimated that 1.27 million deaths were directly attributed to AMR and that 4.95 million deaths were associated with AMR globally in 2019 (1, 2). Beyond its impact on morbidity and mortality, AMR imposes substantial economic consequences on healthcare systems and societies. Studies have shown that antimicrobial resistant infections increase hospital costs by an estimated USD 29,289 per patient and contribute to an estimated global healthcare expenditure exceeding USD 693 billion annually (3, 4). Consequently, AMR is recognised as both a public health emergency and a major threat to sustainable healthcare financing in the world.

The burden of AMR is disproportionately concentrated in low- and middle-income countries (LMICs), particularly those in sub-Saharan Africa (SSA), where infectious diseases remain highly prevalent and antimicrobial medicines are indispensable for clinical care (5). In 2019, an estimated 250,000 deaths were directly attributed to AMR, and 1.05 million deaths were associated with AMR in SSA (6). Lower respiratory tract infection remains among the leading causes of mortality and morbidity in the region, with pathogens such as *Streptococcus pneumoniae*, *Klebsiella pneumoniae*, *Escherichia coli*, and *Staphylococcus aureus* contributing substantially to the disease burden (6).

Malawi, a SSA country, is not spared from the burden of AMR. In 2021, AMR was directly attributed to 2,520 deaths and was associated with 13,837 deaths nationally (7). Additionally, several studies have reported increasing resistance among clinically important pathogens in the country, signalling a worsening AMR landscape and posing significant threats to effective infection management (8–11). Recognising the magnitude of the challenge, Malawi joined other World Health Assembly member states in adopting the updated WHO Global Action Plan on AMR 2026-2036, which identifies optimisation of antimicrobial use as a central pillar for reducing the emergence and spread of resistance (12). Achieving this objective requires a coordinated One Health approach across the human, animal, and environmental sectors, as fragmented implementation may undermine national and global efforts to contain AMR.

Healthcare delivery in Malawi is provided through both public and private sectors, with the latter comprising for-profit providers, non-government organisations and faith-based healthcare institutions. Faith-based health facilities constitute an integral component of health service provision throughout SSA and frequently serve underserved and rural populations. In Malawi, the Christian Health Association of Malawi (CHAM) is the largest faith-based healthcare provider, operating 194 health facilities across 27 of the country’s 28 districts and contributing approximately 37% of national healthcare services (13). Importantly, CHAM facilities serve nearly 75% of remote rural and hard-to-reach populations (14), thereby playing a critical role in achieving the recommended 5–8 km geographical accessibility target for healthcare services. Through its commitment to national health policies and service level agreements with the Government of Malawi, CHAM subsidises maternal, neonatal and child health services to improve access to care. However, resource constraints, supply chain challenges, and parallel health systems may hinder implementation of antimicrobial stewardship (AMS) and antimicrobial use monitoring. Understanding antimicrobial use practices within faith-based health facilities is therefore essential to ensure that national AMR strategies effectively reach all healthcare providers and populations.

Assessment of antimicrobial use is a fundamental component of AMS programmes and a key strategy for combating AMR. To facilitate standardised monitoring, WHO developed drug use indicators, revised in 2006, that evaluate healthcare facility characteristics, prescribing practices, and patient care influencing antimicrobial use (15). Numerous studies globally have assessed antimicrobial use using WHO indicators, and other antimicrobial stewardship metrics; however, most have focused on public healthcare facilities, outpatient settings, specific infectious diseases, or selected prescribing indicators (16–33). Evidence evaluating antimicrobial use comprehensively across healthcare facility, prescribing, and patient-care indicators remains limited, particularly in faith-based healthcare settings. Furthermore, data on antimicrobial use patterns in faith-based health facilities using WHO antimicrobial use indicators are scarce globally, particularly in sub-Saharan Africa, including Malawi.

This study assessed antimicrobial use in CHAM health facilities in Malawi using WHO antimicrobial use indicators. By generating evidence from under-represented but strategically important healthcare sectors, this study provides a case example for faith-based healthcare systems in SSA and contributes to strengthening AMS efforts under Malawi’s national AMR programme and action plan.

## Material and methods

Full details of the study design, sampling procedure, and data collection methods are described in the published study protocol (34); key elements are summarised below.

### Study design and setting

This was a multi-centre cross-sectional study conducted in 29 CHAM health facilities across 13 of the 27 districts where CHAM health facilities are located, covering all three geographical regions of Malawi (Northern, Central and Southern) **S1 Table**. The study assessed antimicrobial use using three WHO antimicrobial use core indicators: health facility, prescribing and patient care indicators (15) **Table 1**. Minimum sample sizes for health facility and prescribing indicators followed WHO recommendations for assessing antimicrobial use in health facilities (15), while the sample size for patient-care indicators was calculated to achieve a pre-specified level of precision (95% CI, ±5%) described in the protocol (34). The study was conducted between January 2024 and June 2025.

**Table 1:**
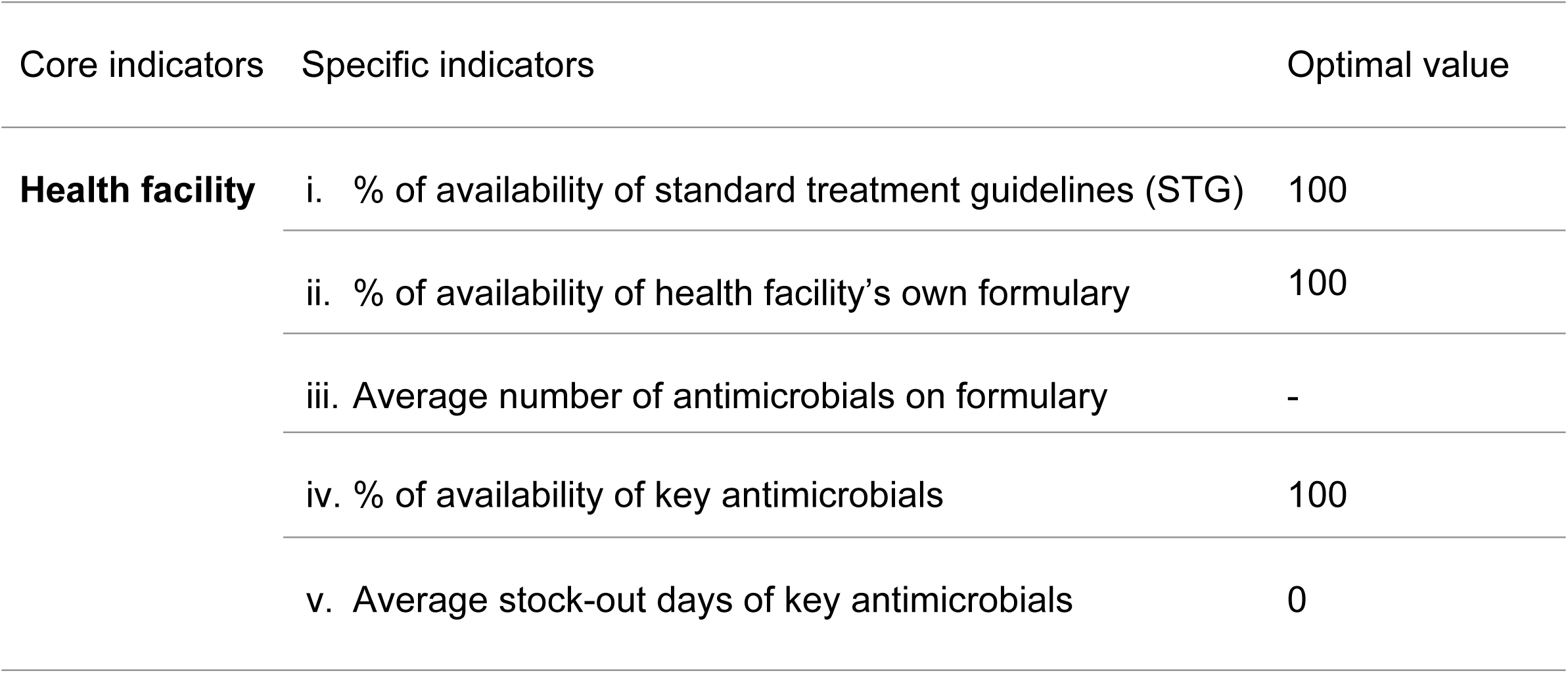
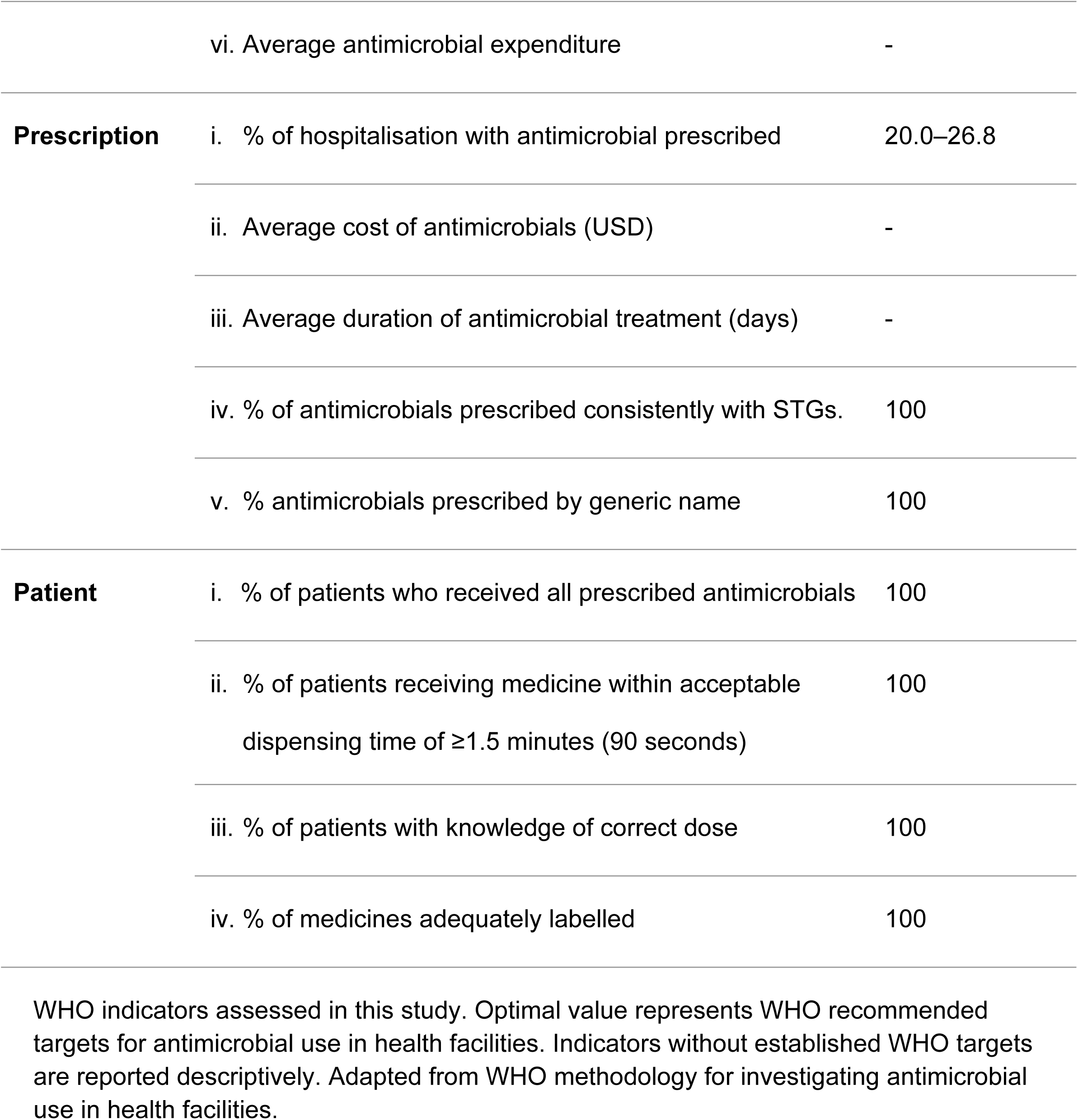
WHO indicators for assessing antimicrobial use in health facilities.

### Study participants

For health facility indicators, randomly selected health facilities that were operational on the day of data collection were included. Health facilities which were temporarily closed because of community-related disputes or lacked admitted patients at the time of data collection, a common occurrence in some CHAM facilities because of their cost-recovery health care service model were replaced through simple random sampling **S1 Table**. Respondents for the health facility indicators were health facility in-charges, including medical directors (medical doctors), clinical in-charges (clinical officers with a diploma in clinical medicine or clinical assistants with a certificate in clinical medicine), nurses, human resource managers, and public administrators (including Catholic nuns and priests). As for prescribing and patient care indicators, inpatient prescriptions with the corresponding patient available for interview were included, while illegible, incomplete, duplicate, or invalid prescriptions were excluded. Children under five years of age were excluded to improve the comparability of prescribing and patient care indicators, as antimicrobial prescribing practices, formulations, and dosing in this age group differ substantially from those in older children and adults.

### Data collection

Data were collected using KoboToolbox v2021.2.4 (35) through standardised electronic questionnaires. Close monitoring ensured data quality and consistency throughout data collection. Each participating health facility appointed a coordinator who received standardised training on prescribing and patient care data collection. Standardised coordinator training and the use of a structured KoboToolBox data-capture form were intended to limit measurement and data-entry bias, while random replacement of temporarily closed or non-functional facilities (described above) was intended to limit selection bias arising from facility substitution. Potential recall or social-desirability bias in patient-reported responses could not be independently verified against an external reference standard. As shown in **Fig 1**, data collection started with communication and courtesy visits to facility in-charges, followed by engagement with ward, pharmacy and data personnel. Data were collected between July 2024 and June 2025.

**Fig 1.**
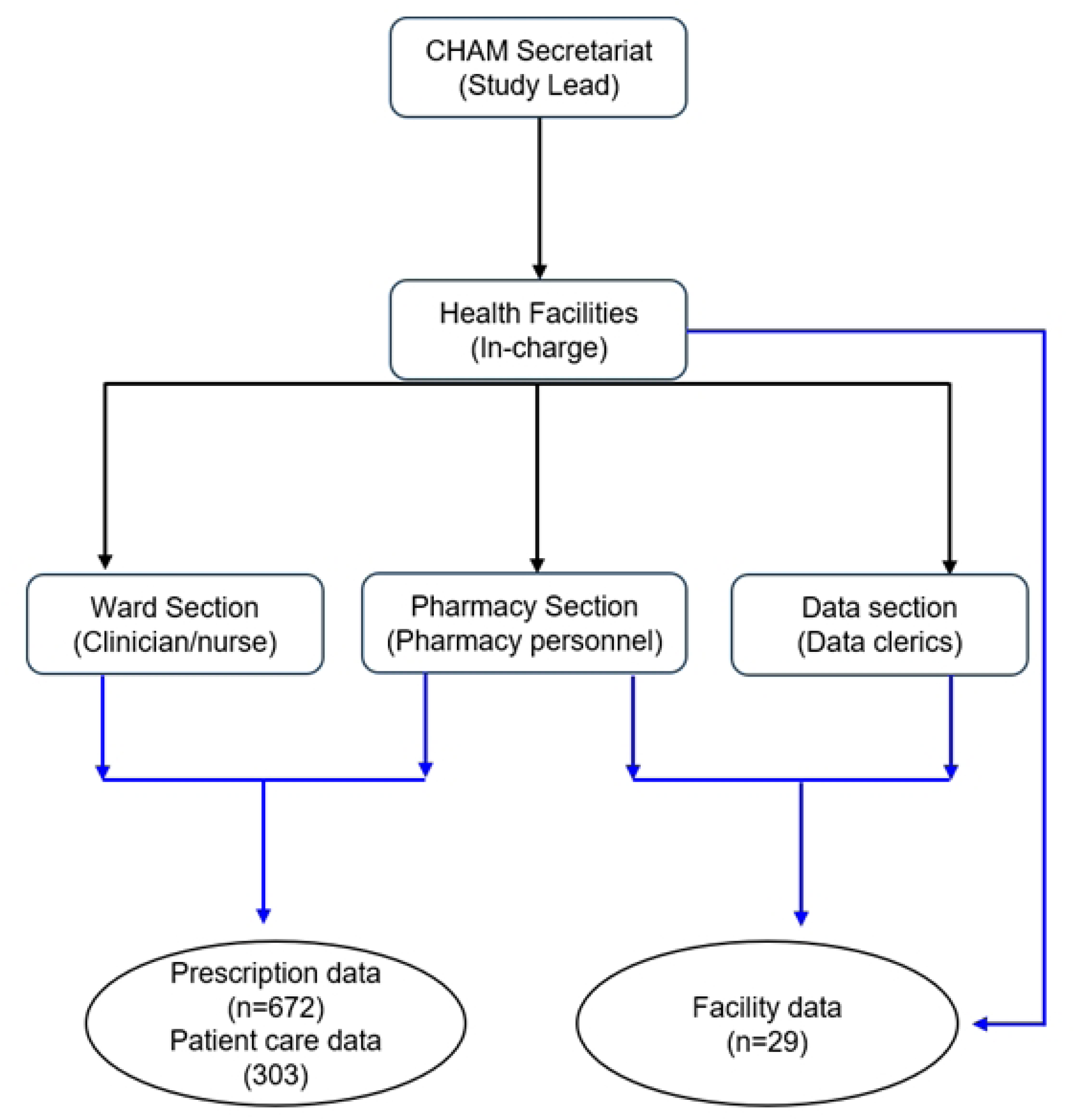
Data collection workflow for the study. Black arrows show communication pathways and movement of the data collection team, while blue arrows indicate data sources. Boxes represent study entities and circles represent data collected.

#### Health facility indicators

Health facility indicators data were collected from 29 randomly selected health facilities in **S1 Table**. Data were obtained through on-site visits or remotely. Clinical in-charges provided information on STGs, pharmacy in-charges provided formulary and antimicrobial availability data, and facility in-charges, accounts personnel, and data clerks provided expenditure, catchment population, and facility characteristic data.

#### Prescribing and patient care indicators

A total of 682 prescriptions were reviewed in accordance with WHO recommendations (15). The protocol specified a target of 22 health facilities, with approximately 30 prescriptions sampled per facility (660 prescriptions in total) (46). Of the 682 prescriptions reviewed, 670 were retained for the final analysis after excluding illegible, incomplete, duplicate, or invalid prescriptions (34). These 670 prescriptions formed the dataset for the prescribing indicators and were also used to assess patient care indicators because they contained all variables required for both assessments, replacing the initially planned sample of 303 participants for the patient care indicators. Data were collected from clinicians, nurses, pharmacy personnel, and patients **Fig 1**.

### Ethical considerations

The study was approved by College of Medicine Research Ethics Committee (COMREC; now Kamuzu University of Health Science Research Ethics Committee [KUREC]) under approval number P.04/24-0651 in May 2024. The approval to conduct the study in CHAM health facilities was subsequently obtained from the CHAM Secretariat management. The study was conducted in accordance with the Declaration of Helsinki. Oral informed consent was obtained from all participants prior to data collection, with assent obtained for participants younger than 18 years. Participants were informed that refusal to participate or withdrawal from the study would not affect the healthcare services they received.

### Data analysis

Data captured using KoboToolBox were downloaded into Microsoft Excel 2016, checked for missing entries, cleaned, and coded. A clinician on the study team created five additional variables: receipt of antimicrobials, number of prescribed antimicrobials per participant, treatment cost based on prevailing Malawian market prices (United States Dollars, USD) (36), treatment duration, and adherence to STGs. Statistical analysis was performed using Stata SE version 14 (37). Descriptive statistics were used to summarise the data, with continuous variables presented as mean (95% confidence interval, CI) or median (interquartile range, IQR) and categorical variables as frequencies and percentages. To assess differences across levels of care, outcomes were stratified by CHAM facility type: full-fledged hospitals (secondary healthcare facilities), community hospitals (intermediate-level facilities), and health centres (primary healthcare facilities).

Associations between categorical variables were assessed using Pearson’s chi-square test, differences in means across facility types were assessed using analysis of variance (ANOVA) or the t-test, as appropriate. Correlations between continuous or ordinal variables were assessed using Spearman’s rank correlation (rho), using a significance level (α) of 0.05. All reported tests were unadjusted bivariate analyses; no multivariable adjustment for potential confounders was performed. Analyses used all available records for each variable (available-case analysis); no imputation was performed for missing data, and denominators are reported alongside each estimate to indicate the amount of missing data. Facilities were sampled as clusters containing multiple prescriptions and patients, but standard errors were not adjusted for facility-level clustering; observations were treated as independent. No additional sensitivity analyses were performed.

## Results

### Baseline characteristics of the study health facilities, prescriptions and patients

A total of 29 CHAM health facilities and 670 prescribing and patient-care records were included in the analysis. Most facilities were health centres (19/29, 65.2%), and consistent with CHAM’s mission of providing healthcare services to underserved populations, nearly half were in hard-to-reach areas (13/28, 46.3%) **Table 2**. Among patients, 396 (59.1%) were female, while one participant preferred not to disclose their sex.

**Table 2:**
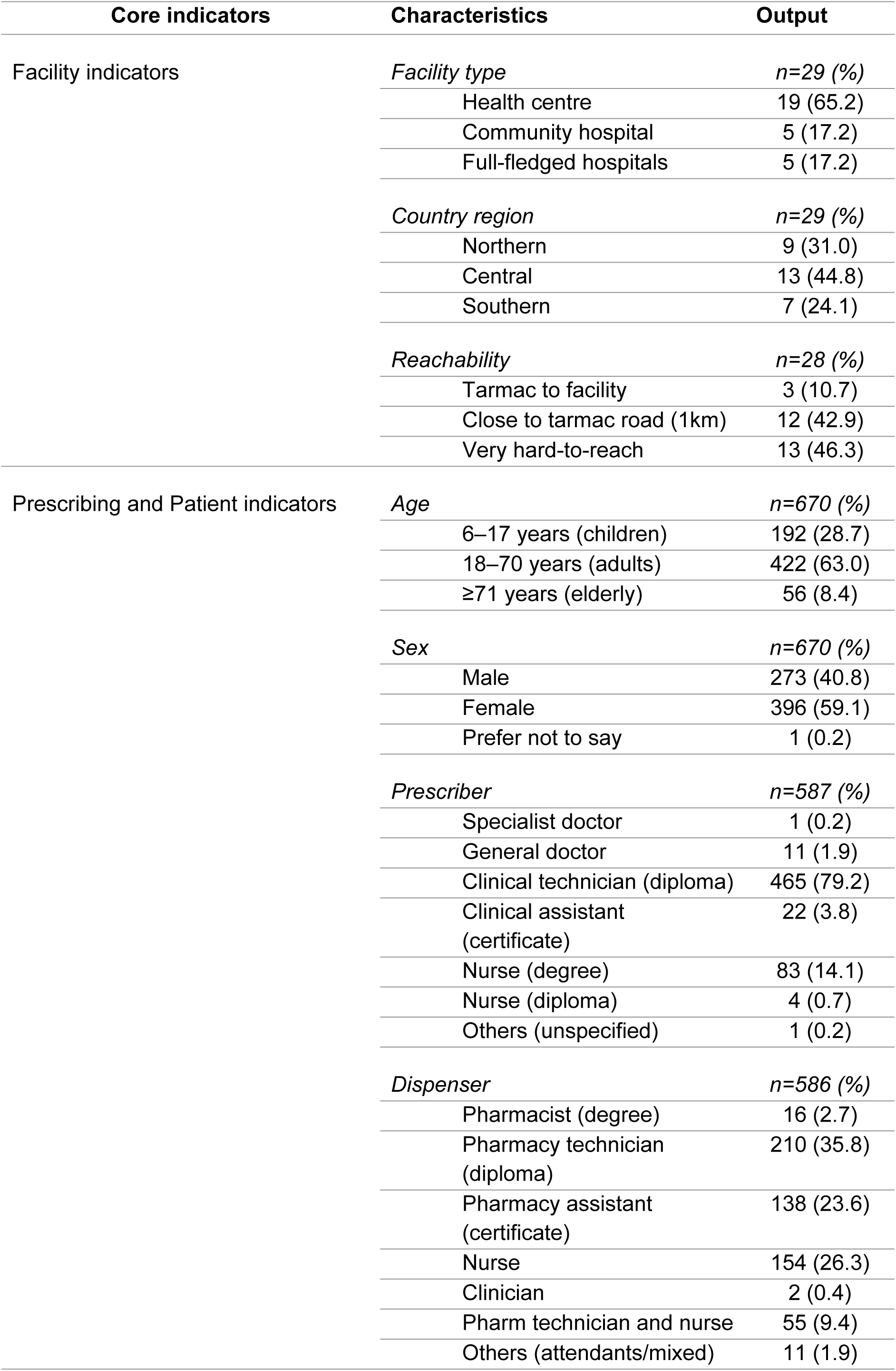
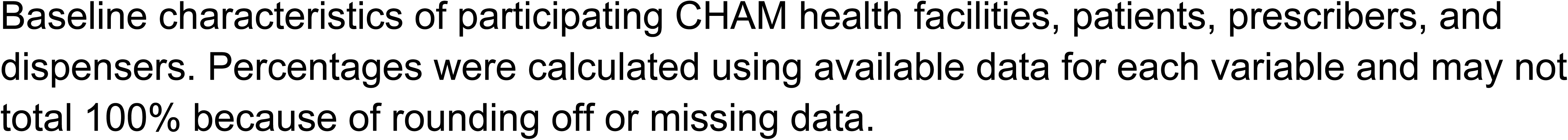
Baseline characteristics of the study.

| Core indicators | Characteristics | Output |
| --- | --- | --- |
| Facility indicators | *Facility type* | *n=29 (%)* |
|  | Health centre | 19 (65.2) |
|  | Community hospital | 5 (17.2) |
|  | Full-fledged hospitals | 5 (17.2) |
|  | *Country region* | *n=29 (%)* |
|  | Northern | 9 (31.0) |
|  | Central | 13 (44.8) |
|  | Southern | 7 (24.1) |
|  | *Reachability* | *n=28 (%)* |
|  | Tarmac to facility | 3 (10.7) |
|  | Close to tarmac road (1km) | 12 (42.9) |
|  | Very hard-to-reach | 13 (46.3) |
| Prescribing and Patient indicators | *Age* | *n=670 (%)* |
|  | 6–17 years (children) | 192 (28.7) |
|  | 18–70 years (adults) | 422 (63.0) |
|  | ≥71 years (elderly) | 56 (8.4) |
|  | *Sex* | *n=670 (%)* |
|  | Male | 273 (40.8) |
|  | Female | 396 (59.1) |
|  | Prefer not to say | 1 (0.2) |
|  | *Prescriber* | *n=587 (%)* |
|  | Specialist doctor | 1 (0.2) |
|  | General doctor | 11 (1.9) |
|  | Clinical technician (diploma) | 465 (79.2) |
|  | Clinical assistant (certificate) | 22 (3.8) |
|  | Nurse (degree) | 83 (14.1) |
|  | Nurse (diploma) | 4 (0.7) |
|  | Others (unspecified) | 1 (0.2) |
|  | *Dispenser* | *n=586 (%)* |
|  | Pharmacist (degree) | 16 (2.7) |
|  | Pharmacy technician (diploma) | 210 (35.8) |
|  | Pharmacy assistant (certificate) | 138 (23.6) |
|  | Nurse | 154 (26.3) |
|  | Clinician | 2 (0.4) |
|  | Pharm technician and nurse | 55 (9.4) |
|  | Others (attendants/mixed) | 11 (1.9) |

Baseline characteristics of participating CHAM health facilities, patients, prescribers, and dispensers. Percentages were calculated using available data for each variable and may not total 100% because of rounding off or missing data.

### Facility indicators

All 29 health facilities responded to the availability of STGs, with 24 (85.7%) reporting availability. Of the 28 facilities that responded to the question on having a facility-specific formulary, 18 (64.3%) had one available **Table 3**. The mean availability of key antimicrobials was 33.1% (95% CI: 29.7–36.4), while the average stock-out duration was 1.4 days (95% CI: 0.77–2.12). Antimicrobials accounted for a median of 40.0% (IQR: 25.3–57.1) of the health facility medicine procurement expenditure.

**Table 3:** Health facility indicators assessment.

| Health facility indicators | Outcome | Optimal value |
| --- | --- | --- |
| Availability of STG (n=29) | 24 (85.7%) | 100 |
| Availability of the facility's own formulary (n=28) | 18 (64.3%) | 100 |
| Average number of antimicrobials on formulary | 30.7 | - |
| Availability of key antimicrobials, % (n=28) | 33.1 (95% CI: 29.7–36.4) | 100 |
| Average key antimicrobials stock-out days (n=27) | 1.4 (95% CI: 0.77–2.12) | 0 |
| Median antimicrobial expenditure, % (n=25)<sup>a</sup> | 40.0 (IQR: 25.3–57.1) | - |

Health facilities indicators where percentages were calculated using available responses for each variable. Denominators may vary because of missing data.
<sup>a</sup>Expenditure was converted from Malawi Kwacha to United States Dollars (USD) using the prevailing Reserve Bank of Malawi exchange rate in June 2025 (36).

The presence of a pharmacy professional at a health facility was significantly associated with the availability of a facility-customised formulary (χ² = 4.37, p = 0.037). Facilities with a pharmacy professional were more likely to have their own formulary than facilities without a pharmacy professional (84.6% vs. 46.7%) **Fig 2**. We found no association between facility type and the availability of key antimicrobials (F[2,26] = 1.65, p = 0.211), the presence of a pharmacy professional and the availability of key antimicrobials at health facilities (t = 1.19, p = 0.245) or the facility reachability and availability of key antimicrobials (F[2,25] = 0.12, p = 0.892).

**Fig 2.**
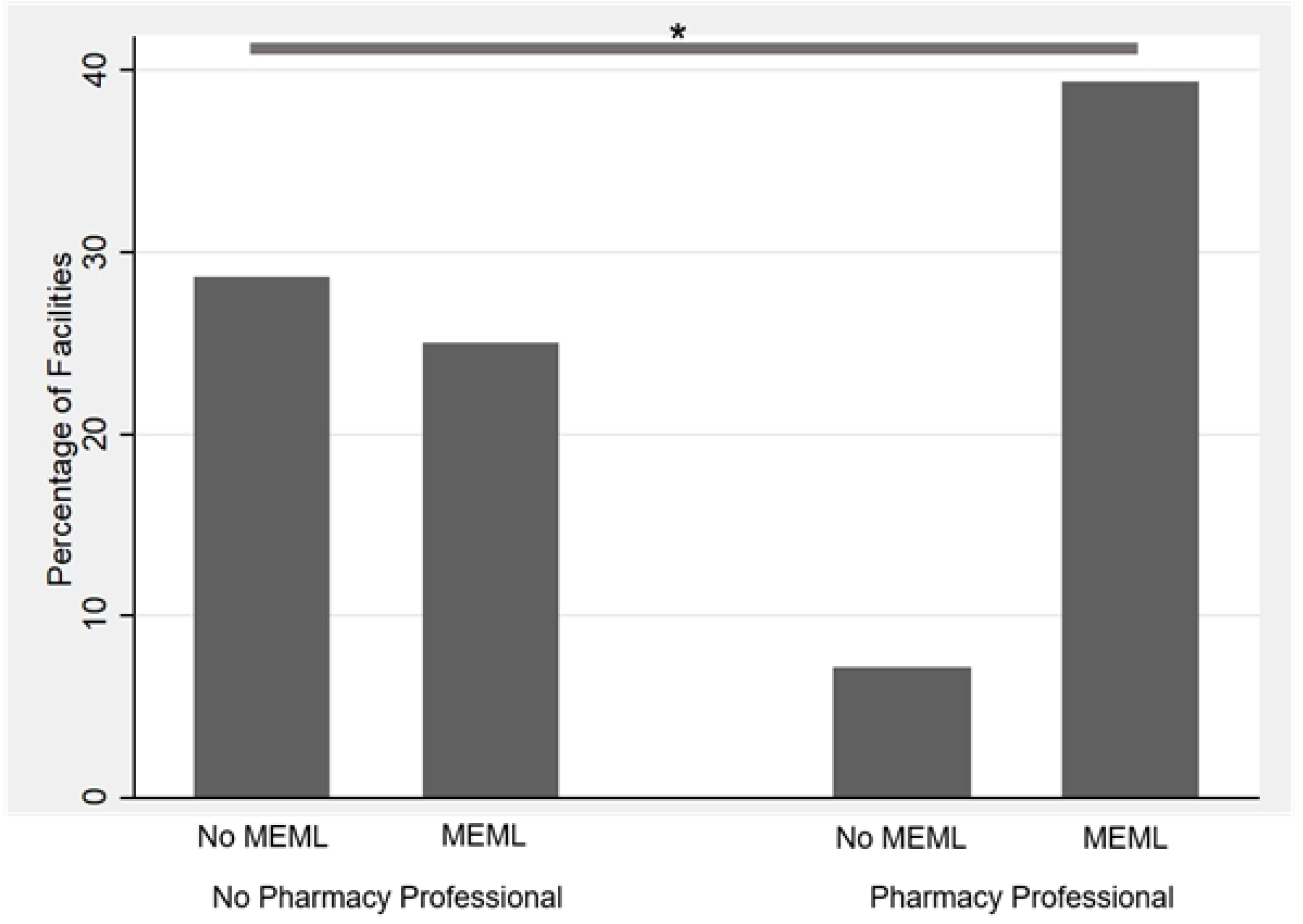
Association between the presence of pharmacy professionals and the availability of an in-house formulary. Pearson’s chi-square test was used to assess the association, with an asterisk (*) indicating a statistically significant association (p < 0.05).

### Prescribing and patient care indicators

For prescribing indicators, 219 (33.2%) antimicrobials were prescribed in accordance with STG, with an average treatment duration of 4.7 days **Table 4**. The average antimicrobial treatment cost was USD 9.1, while generic prescribing was 43.6%; most prescriptions (1,205; 53.9%) used abbreviated generic names **S2 Fig**. For patient care indicators, 568 (98.1%) patients received all prescribed antimicrobials. Responses were available for 45 and 47 patients on knowledge of the correct dose and medicine labelling, respectively. Of these, 38 (84.4%) correctly stated the dose of their treatment, while only 17 (36.2%) received adequately labelled medicines.

**Table 4:**
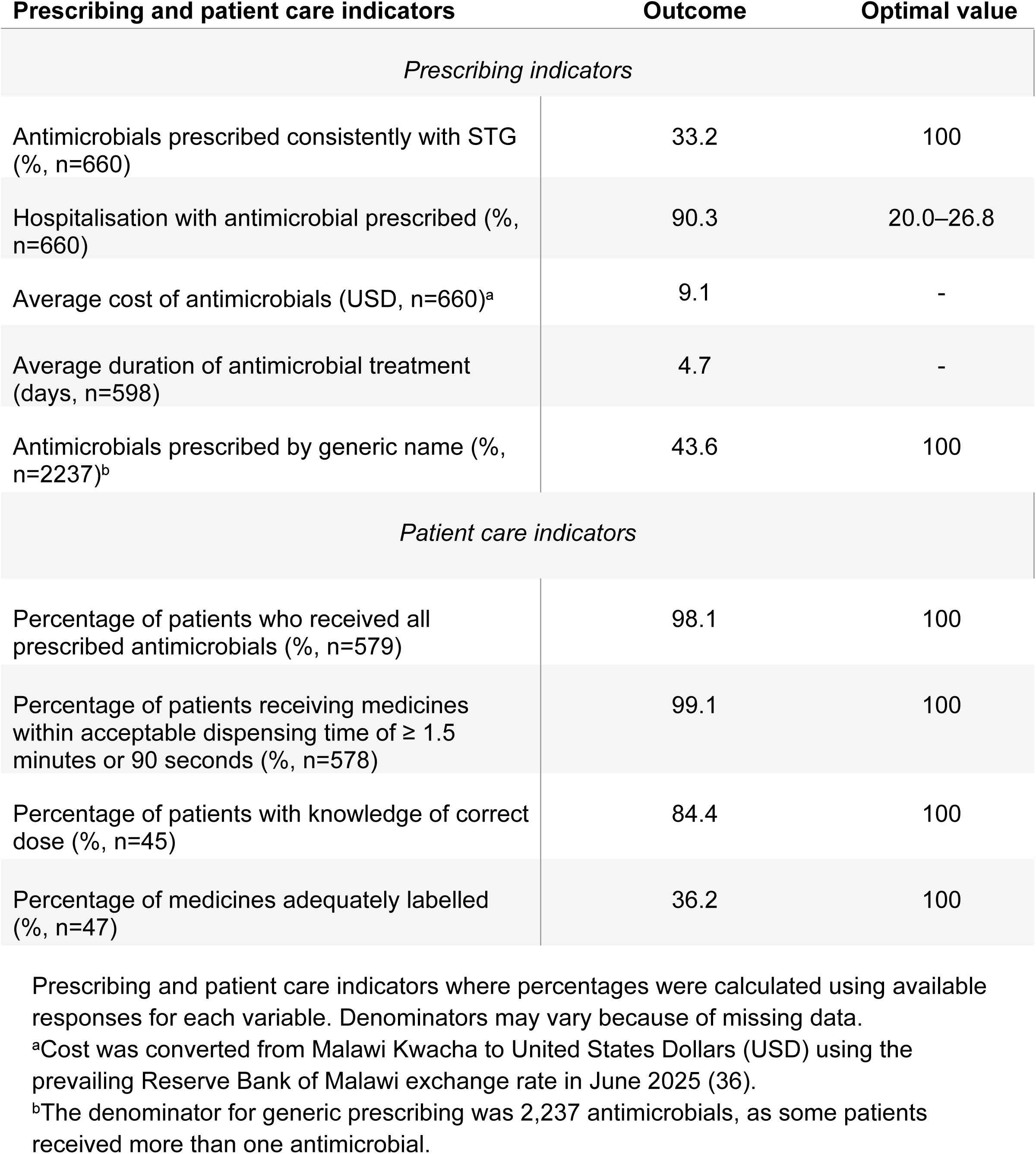
Assessment of prescribing and patient care indicators.

Patients attending community hospitals were significantly more likely to receive antimicrobial treatment than those attending health centres or full-fledged hospitals (χ² = 13.24, p = 0.001) **Fig 3**. There was also a significant association between patient age and receipt of an antimicrobial prescription (χ² = 29.95, p < 0.001), with the likelihood of receiving an antimicrobial increasing with age **Fig 4**. The number of prescribed antimicrobials was positively correlated with antimicrobial treatment cost (Spearman’s rho = 0.776, p < 0.001) **S2 Fig**. Prescriber professional level did not significantly influence antimicrobial prescribing (F(6,573) = 1.75, p = 0.107).

**Fig 3.**
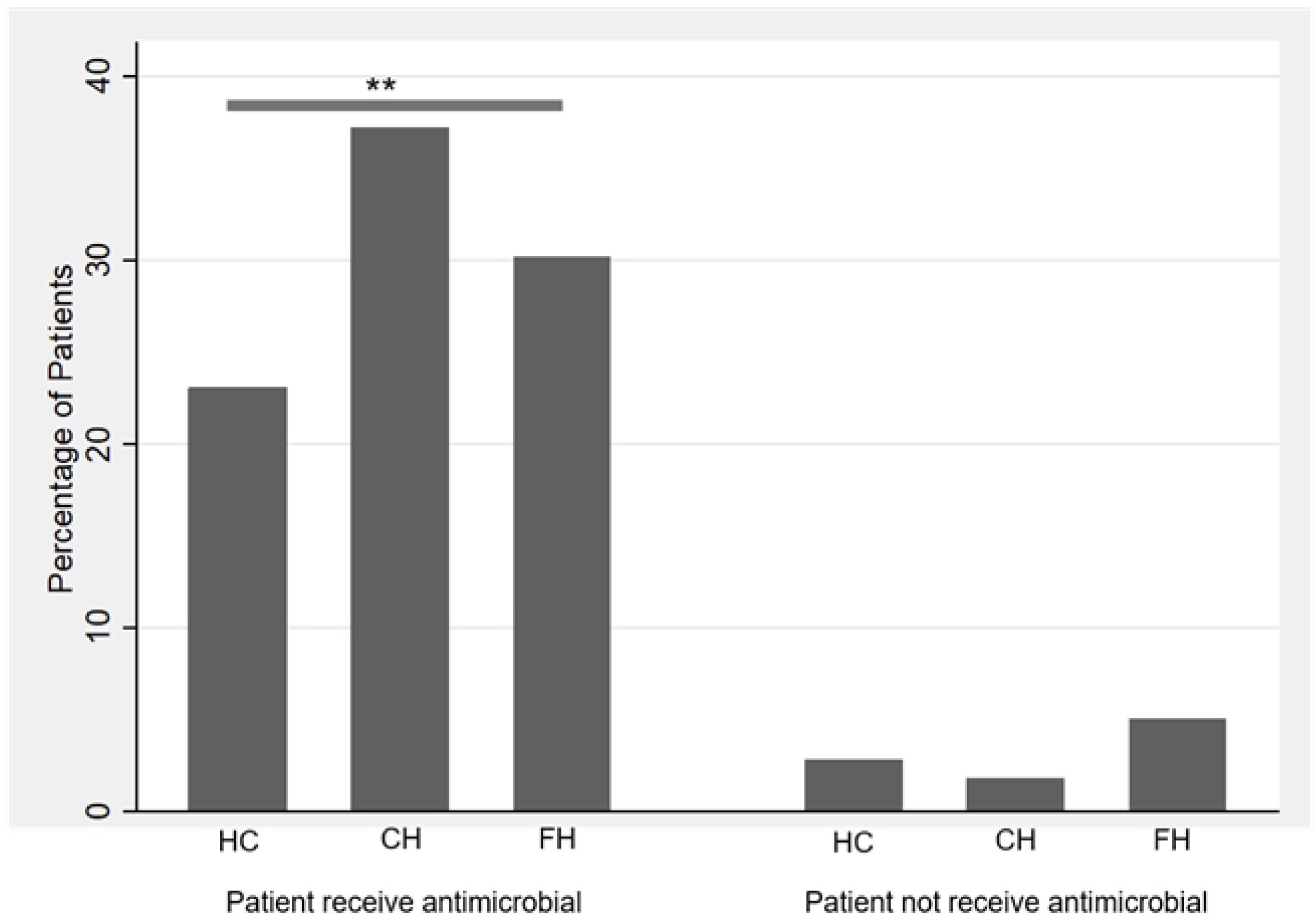
Association between the facility type and patients receiving antimicrobial treatment. Pearson’s chi-square test was used to assess the association, with an asterisk (*) indicating a statistically significant association (p < 0.05).

**Fig 4.**
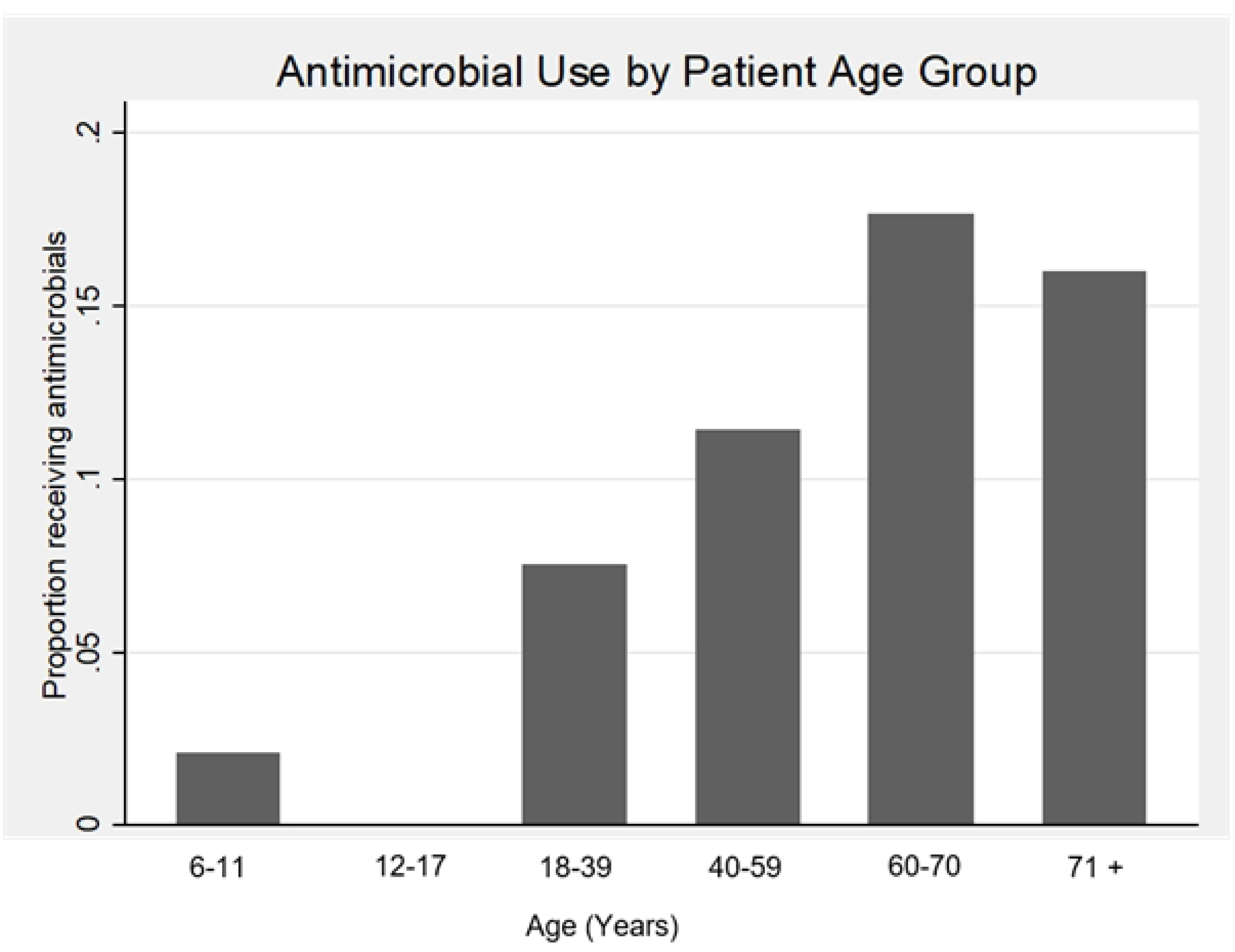
Association between patient age and receipt of an antimicrobial prescription. Asterisks (*) indicate statistically significant associations based on Pearson’s chi-square test (p < 0.05).

## Discussion

This study provides the very first comprehensive assessment of antimicrobial use using the three WHO core indicators within faith-based health facilities in a sub-Saharan African country. The findings demonstrate significant gaps in AMS implementation across CHAM health facilities despite their major contribution to healthcare delivery in Malawi, particularly for rural and underserved populations, who comprise more than 80% of the country’s population (38). At the health facility level, availability of STG (85.7%) and facility customised formularies (64.3%) remained below the WHO target of 100%, suggesting incomplete stewardship infrastructure. Similar gaps have been reported in hospitals in Zambia (31, 39). Importantly, health facilities with pharmacy professionals were significantly more likely to possess facility-specific formularies, reinforcing evidence that pharmacists play a critical role in AMS through formulary management, guideline implementation, clinical audit, and optimisation of prescribing practices (40, 41). However, the most concerning health facility finding was the low availability of key antimicrobials (33.1%), which was considerably lower than values reported from Palestinian hospitals and other healthcare settings (26). Although average stock-out duration was relatively short, poor antimicrobial availability may compel prescribers to select medicines based on availability rather than clinical appropriateness, potentially increasing inappropriate use of broad-spectrum agents and accelerating AMR. Furthermore, antimicrobials accounted for approximately 40% of medicine procurement expenditure, highlighting their substantial financial burden on faith-based healthcare systems. Similar observations have been reported globally, where antimicrobial procurement represents a major component of healthcare expenditure and contributes substantially to the economic burden associated with AMR (2, 42). These findings collectively suggest that strengthening pharmaceutical supply systems and pharmacy workforce capacity should be central components of antimicrobial stewardship programmes within faith-based health facilities.

The prescribing indicators revealed substantial departures from WHO recommendations, suggesting opportunities for stewardship improvement. The prevalence of antimicrobial prescribing among hospitalised patients was 90.3%, which greatly exceeds the WHO optimal range of 20.0–26.8% and is considerably higher than reports from Zambia (59.0%), Afghanistan (49.1%), Tanzania (26.1%), Saudi Arabia (26.5%), and Ghana (18–36%) (21–23, 25, 27). This high prevalence likely reflects the substantial burden of infectious diseases, reliance on empirical treatment, limited access to microbiological diagnostics, and the inpatient nature of the study population. Nevertheless, such extensive antimicrobial exposure creates substantial selective pressure for resistance development. Equally concerning was the low adherence to STGs, with only one-third of antimicrobial prescriptions consistent with recommended treatment protocols. Although this finding was slightly higher than that reported in Zambian hospitals (27%), it remains substantially below levels reported in facilities with more established AMS programmes (27, 43). Poor adherence may reflect medicine availability constraints, limited access to updated treatment guidelines, inadequate stewardship oversight, and diagnostic uncertainty. Generic prescribing was also low at 43.6%, falling below values reported from Afghanistan, Ghana, Pakistan, and Yemen (20–22). However, more than half of prescriptions used abbreviated generic names, suggesting that generic prescribing itself was common, but documentation practices were suboptimal. While abbreviated names may improve efficiency, they increase the risk of medication errors, misinterpretation, and dispensing inaccuracies. The average antimicrobial treatment duration of 4.7 days is consistent with the increasing global shift towards shorter treatment courses where clinically appropriate, although future studies should evaluate indication-specific treatment durations to determine whether observed prescribing patterns align with evidence-based recommendations.

Patient care indicators presented a mixed picture. On one hand, almost all patients received their prescribed antimicrobials (98.1%), and nearly all medicines were dispensed within the WHO-recommended dispensing time (99.1%). These findings are encouraging and likely reflect the CHAM cost-recovery healthcare model, where medicines are procured and available before treatment is initiated. Unlike many public sector facilities affected by medicine shortages, patients attending CHAM health facilities may therefore have improved access to prescribed antimicrobials. However, the possibility that clinicians prescribe according to medicine availability rather than optimal therapeutic choice cannot be excluded, particularly given the limited availability of key antimicrobials identified in this study. This concern becomes even more important when interpreted alongside evidence from AMS literature showing that medicine shortages frequently alter prescribing behaviour and may increase use of inappropriate alternatives (44, 45). While access to prescribed antimicrobials was high, patient understanding of treatment and dispensing quality remained suboptimal. Only 84.4% of patients correctly understood their dosage instructions, and merely 36.2% received adequately labelled medicines. Similar deficiencies have been reported across several low- and middle-income countries and are recognised contributors to poor adherence, medication errors, treatment failure, and inappropriate antimicrobial use (46–48). Strengthening pharmacy services, improving dispensing practices, implementing standardised labelling procedures, and enhancing patient counselling therefore represent important steps for improving both AMS and patient outcomes within faith-based healthcare settings.

The study also identified several factors associated with antimicrobial use. Patients attending community hospitals were significantly more likely to receive antimicrobials than those attending health centres or full-fledged hospitals, suggesting that facility-level characteristics, case mix, and prescribing cultures influence antimicrobial utilisation. Similarly, increasing age was associated with a higher likelihood of receiving antimicrobials, which is consistent with evidence showing greater infection burden, increased comorbidity, and more frequent healthcare contact among older patients (49, 50). In contrast, prescriber professional level did not significantly influence antimicrobial prescribing practices, suggesting that institutional factors may exert a stronger influence on prescribing behaviour than individual professional training alone. This observation aligns with stewardship literature demonstrating that prescribing behaviour is often shaped by organisational culture, medicine availability, treatment guidelines, supervision structures, and healthcare system constraints rather than professional cadre alone (51).

This study has several limitations. Its cross-sectional design limits causal inference, and findings from CHAM facilities may not be fully generalisable to public or private healthcare settings. Patient knowledge and medicine-labelling indicators were based on relatively few respondents because of missing data. The number of prescriptions analysed (670) differs from the number planned in the protocol (660) and the number reviewed (682); although this discrepancy is reconciled in the Methods, any unmeasured difference between excluded and included prescriptions could introduce a degree of selection bias or imprecision not fully captured by the reported confidence intervals. All associations reported in this study (chi-square, ANOVA/t-test, Spearman’s rho) are unadjusted bivariate estimates; because facilities were sampled as clusters of multiple prescriptions and patients, but clustering was not accounted for in the analysis, standard errors and confidence intervals may be underestimated and statistical significance overstated. Nevertheless, this study provides important evidence on AMS in faith-based health facilities in Malawi and, through its nationally representative geographical and facility-level coverage, offers findings that may inform AMS strengthening in similar faith-based healthcare settings across sub-Saharan Africa.

## Conclusion

This study demonstrates that AMS challenges remain substantial within faith-based health facilities in Malawi, particularly in prescribing practices, guideline adherence, and antimicrobial availability. Given the important role of faith-based providers in healthcare delivery across sub-Saharan Africa, these findings highlight critical opportunities to strengthen antimicrobial use and support AMR containment. Future studies should evaluate the clinical appropriateness of antimicrobial prescribing using microbiological investigations, culture-guided therapy, antimicrobial resistance patterns, and patient outcomes to better inform stewardship interventions.

## Data Availability

All relevant data underlying the findings of this study are provided within the manuscript and its Supporting Information files. For Excel files, key is in the header of analysed data.

## Author’s contribution

All authors made substantial contributions to the conception, study design, data acquisition, analysis, interpretation, and execution of the study; participated in drafting, revising, or critically reviewing the manuscript; approved the final version for publication; agreed on the target journal; and accepted responsibility for all aspects of the work.

## Acknowledgement

The authors acknowledge COMREC (now KUREC) for reviewing and approving the study protocol, and the CHAM Secretariat for administrative and financial support. Appreciation is also extended to the management of all participating health facilities for their cooperation and support. Special recognition goes to the data collecting team who dedicated their time to this study: Reuben Kachingwe, Sister Constance Moyonsana, Alepher Mphero, Blessings Ndale, Happiness Jolie Makuluni, Haswell Mwale, Ishmael Jimu, Joel Mbene, Keniace Damalekani, Kelita Chizinga, Martha Magalasi, Massa Grafar Mkwende, Moses Maudzu, Dominique Mhango, Samson Mwango, Steven Patrick Msusa, Thembie Nkhoma, Victor Msiska, Grace Mwale, Chimwemwe Mzumala, James Edward Kamanga, Grant Kanje, Catherine Sunde, Constance Chaweka, Brian Mdhuli, Mc Donald Ngulube, Tionge Madula, and Kondwani Kumwenda.

## Supporting information

**S1 Fig. Classification of antimicrobial prescribing by naming convention.** Denominators represent the total number of antimicrobials prescribed rather than the number of patients, as some patients received more than one antimicrobial. Percentage represents what the number represents out of overall medicines prescribed.

**S2 Fig. Correlation between the number of prescribed antimicrobials and antimicrobial treatment cost**. Each point represents an individual patient prescription, while the Locally Weighted Scatterplot Smoothing curve illustrating the overall trend.

**S1 Table: Health facilities, prescriptions and patients in the study**. The health facilities included those replacing closed or not reachable health facilities. For each facility, number of prescriptions is indicated.

**S2 Table: Health facilities dataset**. The header row of the Excel file contains the variable names and definitions (key) for all analysed variables.

**S3 Table: Prescription and patient dataset**. The header row of the Excel file contains the variable names and definitions (key) for all analysed variables.

